# The Bedtime Trap: Smartphone Use Until Sleep Onset and Its Association With Sleep Quality and Academic Performance Among Medical Students in Punjab, Pakistan: A Cross-Sectional Survey

**DOI:** 10.64898/2026.05.30.26354530

**Authors:** Mobeen Sajjad

## Abstract

**Background:** Smartphone use among medical students has become pervasive. While existing literature links excessive smartphone use to poor sleep quality, the specific behavioral pattern most strongly associated with sleep disruption remains insufficiently characterized. This study investigated whether the timing of smartphone cessation relative to sleep onset is more strongly associated with poor sleep quality than total daily screen time among medical students in Punjab, Pakistan, and examined the moderating role of exam period status.

**Methods:** A cross-sectional anonymous online survey was conducted among medical students across Punjab, Pakistan (May 2026). Sleep quality was assessed using items informed by Pittsburgh Sleep Quality Index (PSQI) response formats [1]. Descriptive statistics, chi-square tests, and binary logistic regression were applied to 369 eligible responses, reported in accordance with STROBE guidelines.

**Results:** Of 369 respondents (49.9% female, 48.2% male), 74.8% reported using smartphones 6 or more hours daily and 61.2% used their smartphone until falling asleep. Overall, 75.7% reported poor sleep quality. Students using smartphones until sleep onset had 95.1% poor sleep quality compared to 44.8% in those who ceased use before sleeping (p less than 0.001). In logistic regression with both variables entered simultaneously, bedtime use until sleep onset remained independently associated with poor sleep quality (OR 15.3, 95% CI 5.7-41.2, p less than 0.001), while total daily screen time lost significance (OR 1.8, 95% CI 0.7-4.7, p=0.228). Outside exam periods, 99.0% of students using smartphones until sleep onset reported poor sleep quality versus 24.2% of those who stopped before sleeping -- a difference of 74.8 percentage points (p less than 0.001). During exam periods, no significant association was observed (p=0.075), suggesting exam-related stress may attenuate the bedtime behavior effect. Hostel-dwelling students showed the highest prevalence of bedtime smartphone use, with 79.0% using smartphones until sleep onset compared to 23.2% of family-living students (p less than 0.001).

**Conclusions:** Bedtime smartphone use until sleep onset is more strongly associated with poor sleep quality than total daily screen time among Pakistani medical students. Medical institutions should consider integrating targeted digital wellness education -- specifically addressing bedtime cessation timing -- into student health programs, with particular attention to hostel-dwelling students.

## 1. Introduction

Smartphone use among medical students has expanded rapidly, driven by the utility of mobile devices for accessing clinical decision support, medical literature, communication platforms, and educational resources [2,3]. Systematic reviews and meta-analyses have consistently documented that this intensive digital engagement is associated with poor sleep quality and impaired academic performance -- two outcomes that are closely and bidirectionally linked in medical student populations [2,3] and broader university student populations [4,5].

The relationship between smartphone use and sleep disruption is well-documented. Proposed mechanisms include blue light exposure suppressing melatonin secretion, cognitive and emotional arousal from social media engagement, and conditioned wakefulness associated with device proximity [6,7]. Studies examining digital device use, including smartphone and internet use, have demonstrated associations with poor sleep quality using validated instruments in medical and health sciences student populations [8,9,10,11,12].

However, a critical gap persists in this literature: most published studies characterize smartphone use through aggregate measures -- total daily screen time or composite addiction scale scores -- without distinguishing between the timing and pattern of use relative to the sleep-wake cycle. This distinction may be behaviorally important, as the physiological and psychological mechanisms linking smartphone use to sleep disruption are likely most active in the period immediately preceding sleep onset [6,7]. Studies examining electronic device use at bedtime have reported associations with shorter sleep duration and poorer sleep quality [11], and objective wearable sensor data specifically confirm that smartphone use in bed -- not outside bed -- significantly worsens sleep latency and awake time [13].

Available evidence suggests that bedtime cessation behavior has not been examined as a primary exposure variable in Pakistani medical student populations. Prior studies from Pakistani medical schools have documented high rates of sleep disruption associated with academic stress [14], and smartphone addiction has been reported as prevalent among Pakistani medical and dental students [15]. Pakistan maintains one of the largest medical education systems in South Asia, and its healthcare system faces significant resource constraints that limit institutional capacity for student wellness programs [16]. This gap represents an opportunity to characterize smartphone-sleep associations in an LMIC setting that is structurally distinct from populations studied in existing literature.

This study was designed to address this gap. We hypothesized that bedtime smartphone cessation behavior would demonstrate a stronger association with sleep quality than total daily screen time, and that this association would be moderated by exam period status and residential setting.

## 2. Methods

### 2.1 Study Design

We conducted a cross-sectional, self-administered, anonymous online survey, designed and reported in accordance with the Strengthening the Reporting of Observational Studies in Epidemiology (STROBE) guidelines for cross-sectional studies.

### 2.2 Setting and Participants

Participants were recruited via WhatsApp medical student networks, LinkedIn, and direct academic contacts across Punjab, Pakistan during May 2026. Eligibility criteria were: currently enrolled as a medical student (years 1-5) in Pakistan, aged 17 years or older, and provision of voluntary informed consent. No specific institutional affiliation was required for eligibility.

### 2.3 Survey Instrument

The survey was administered using Google Forms and comprised 30 items across four domains: (1) demographic and academic characteristics; (2) smartphone use patterns including daily hours, primary purpose, bedtime use, cessation timing, nocturnal checking behavior, and screen time management; (3) sleep characteristics using items informed by Pittsburgh Sleep Quality Index (PSQI) response formats [1], including bedtime, sleep onset latency, total sleep duration, and overall sleep quality rating; and (4) academic performance and related factors including self-reported academic standing, study hours, stress level, and perceived impact of smartphone use and sleep on academic performance.

Sleep quality was operationalized using the overall subjective sleep quality item (Item 6 of the PSQI), which asks respondents to rate their sleep quality overall as very good, fairly good, fairly bad, or very bad [1]. For primary analyses, responses were dichotomized as poor (fairly bad or very bad) or good (fairly good or very good). Sleep onset latency was measured using PSQI-derived categorical response options. The exposure variable of primary interest was the response to: ‘When do you usually stop using your smartphone before sleeping?’ with options ranging from ‘I use my smartphone until I fall asleep’ to ‘I do not use my smartphone before sleeping,’ dichotomized as ‘uses until sleep onset’ versus ‘stops before sleeping.’ This behavioral cessation variable was chosen in preference to a composite smartphone addiction scale, as the research question focused specifically on pre-sleep behavioral patterns rather than addiction construct.

### 2.4 Ethical Considerations

This study involved anonymous, voluntary participation with no collection of personal identifiers, no clinical intervention, and no patient data. All participants provided explicit informed consent via a mandatory confirmation item at the start of the survey. The study qualifies for exemption from formal institutional review board review under standard criteria for anonymous survey research. AI tools were used for language refinement during manuscript preparation. All scientific content, interpretations, and references were developed and independently verified by the author.

### 2.5 Statistical Analysis

Data were exported from Google Forms and analyzed using Python (pandas, scipy, and scikit-learn libraries). Frequencies and percentages were calculated for all categorical variables. Chi-square tests of independence assessed associations between smartphone behavioral variables and sleep quality, sleep onset latency, and academic performance. Stratified analyses by exam period status and residential setting examined potential effect modification. Binary logistic regression was performed with poor sleep quality as the outcome to assess the independent associations of bedtime cessation behavior and total daily screen time when entered simultaneously, and in a fully adjusted model additionally including exam period status, residential setting, and nocturnal phone checking. Odds ratios (OR) with 95% confidence intervals (CI) are reported. A p-value of less than 0.05 was considered statistically significant. Given the exploratory nature of secondary analyses, no corrections for multiple comparisons were applied; secondary results are hypothesis-generating.

## 3. Results

### 3.1 Participant Characteristics

A total of 369 medical students completed the survey. All confirmed current enrollment at a Pakistani medical institution and provided informed consent. No responses were excluded. Participant characteristics are summarized in Table 1.

**Table 1.** Demographic and academic characteristics of survey participants (N=369)

| Characteristic | N | % |
| --- | --- | --- |
| Year of study: 1st year (Pre-clinical) | 35 | 9.5 |
| Year of study: 2nd year (Pre-clinical) | 43 | 11.7 |
| Year of study: 3rd year (Clinical) | 89 | 24.1 |
| Year of study: 4th year (Clinical) | 93 | 25.2 |
| Year of study: Final year (Clinical) | 109 | 29.5 |
| Gender: Female | 184 | 49.9 |
| Gender: Male | 178 | 48.2 |
| Age: 17-19 years | 24 | 6.5 |
| Age: 20-22 years | 92 | 24.9 |
| Age: 23-25 years | 148 | 40.1 |
| Age: 26 years or older | 105 | 28.5 |
| Residential status: Hostel/student accommodation | 214 | 58.0 |
| Residential status: Living with family | 95 | 25.7 |
| Residential status: Private rented accommodation | 60 | 16.3 |
| Currently in exam period: Yes | 141 | 38.2 |
| Currently in exam period: No | 228 | 61.8 |

Final year students comprised the largest group (29.5%, n=109), followed by 4th year (25.2%, n=93) and 3rd year (24.1%, n=89). Pre-clinical students (years 1-2) accounted for 21.2% (n=78). Gender distribution was near-equal (49.9% female, 48.2% male). The majority were aged 23-25 years (40.1%). Hostel or student accommodation was the most common residential setting (58.0%, n=214), followed by living with family (25.7%, n=95) and private rented accommodation (16.3%, n=60). At the time of survey completion, 38.2% (n=141) reported being in an exam period.

### 3.2 Smartphone Use Patterns

Smartphone use was near-universal and intensive. A total of 74.8% (n=276) reported using smartphones for 6 or more hours daily; 57.5% (n=212) reported more than 8 hours of daily use. Social media was the primary use for 36.0% (n=133), followed by academic purposes (31.7%, n=117) and entertainment (23.0%, n=85) -- a pattern consistent with prior reports from Pakistani medical student populations [12,15]. The majority (61.2%, n=226) reported using smartphones in bed every night; the same proportion (61.2%, n=226) reported using their smartphone until falling asleep. A further 15.7% (n=58) stopped less than 30 minutes before sleeping. Only 4.9% (n=18) reported not using their smartphone before sleeping. A total of 62.9% (n=232) always kept their smartphone within reach while sleeping, and 59.9% (n=221) checked their smartphone every night between midnight and wake time -- a pattern consistent with prior evidence linking nocturnal technology use with adverse sleep-related outcomes [17]. The most common bedtime was after 2:00 AM (49.1%, n=181).

### 3.3 Sleep Characteristics

The majority of respondents reported poor sleep quality: 45.3% (n=167) reported very bad sleep quality and 30.4% (n=112) reported fairly bad sleep quality, yielding a combined poor sleep prevalence of 75.7%. Only 5.7% (n=21) reported very good sleep quality. Sleep onset latency was prolonged in the majority: 60.7% (n=224) reported taking more than 60 minutes to fall asleep and a further 26.3% (n=97) reported 31-60 minutes, yielding 87.0% with sleep onset latency greater than 30 minutes -- substantially higher than rates reported in comparable published studies [8,9]. Total sleep duration was 5-6 hours for 57.5% (n=212) and less than 5 hours for 4.9% (n=18).

### 3.4 Bedtime Cessation Behavior and Sleep Quality

Students who used their smartphone until falling asleep had substantially worse sleep quality than those who ceased use before sleeping. Among students using smartphones until sleep onset (n=226), 95.1% reported poor sleep quality, compared to 44.8% of those who stopped before sleeping (n=143) -- a difference of 50.3 percentage points (chi-square=117.814, p less than 0.001) (Table 2).

**Table 2.**
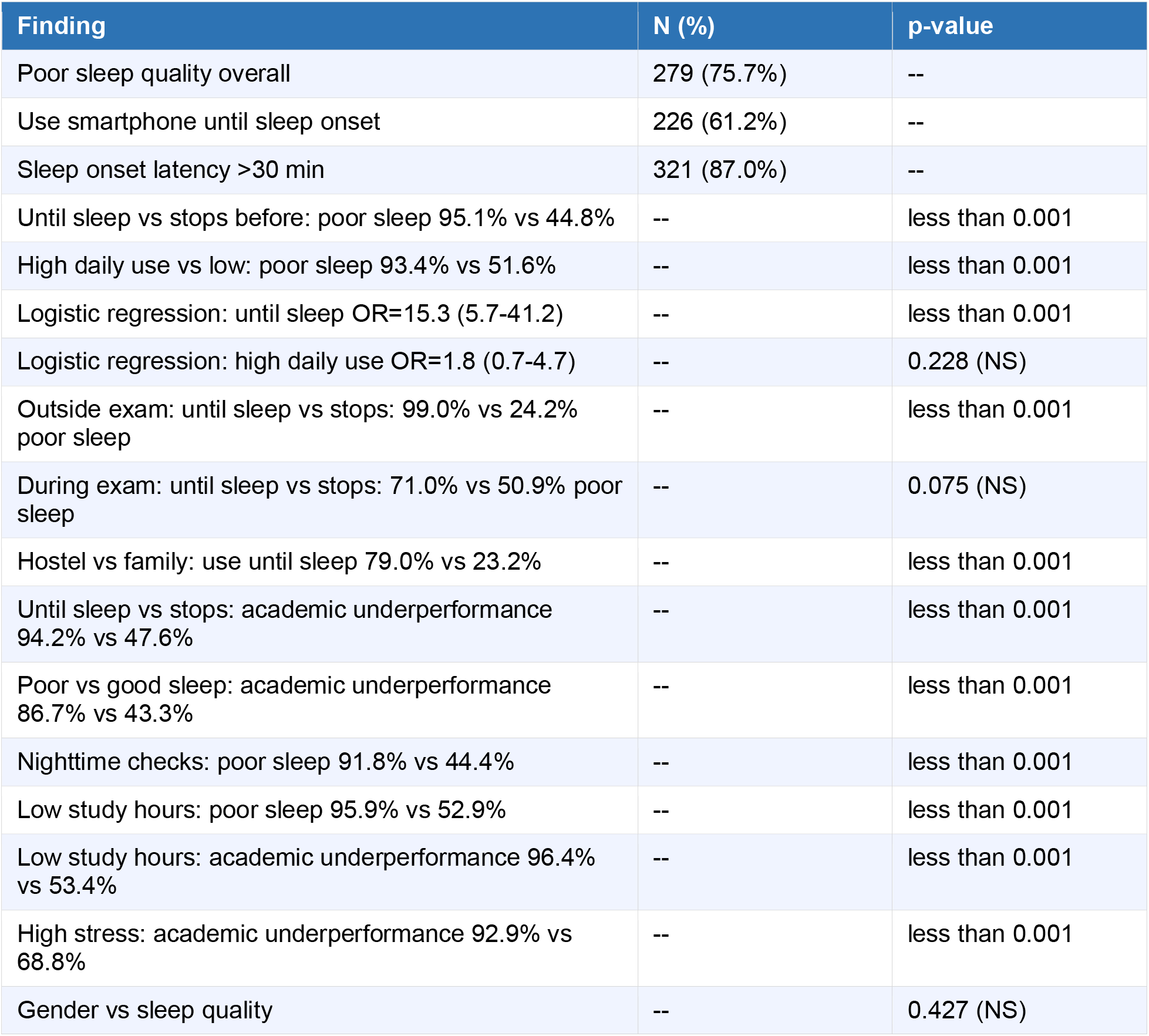
Key findings: smartphone use, sleep quality, and academic performance (N=369)

| Finding | N (%) | p-value |
| --- | --- | --- |
| Poor sleep quality overall | 279 (75.7%) | -- |
| Use smartphone until sleep onset | 226 (61.2%) | -- |
| Sleep onset latency >30 min | 321 (87.0%) | -- |
| Until sleep vs stops before: poor sleep 95.1% vs 44.8% | -- | less than 0.001 |
| High daily use vs low: poor sleep 93.4% vs 51.6% | -- | less than 0.001 |
| Logistic regression: until sleep OR=15.3 (5.7-41.2) | -- | less than 0.001 |
| Logistic regression: high daily use OR=1.8 (0.7-4.7) | -- | 0.228 (NS) |
| Outside exam: until sleep vs stops: 99.0% vs 24.2% poor sleep | -- | less than 0.001 |
| During exam: until sleep vs stops: 71.0% vs 50.9% poor sleep | -- | 0.075 (NS) |
| Hostel vs family: use until sleep 79.0% vs 23.2% | -- | less than 0.001 |
| Until sleep vs stops: academic underperformance 94.2% vs 47.6% | -- | less than 0.001 |
| Poor vs good sleep: academic underperformance 86.7% vs 43.3% | -- | less than 0.001 |
| Nighttime checks: poor sleep 91.8% vs 44.4% | -- | less than 0.001 |
| Low study hours: poor sleep 95.9% vs 52.9% | -- | less than 0.001 |
| Low study hours: academic underperformance 96.4% vs 53.4% | -- | less than 0.001 |
| High stress: academic underperformance 92.9% vs 68.8% | -- | less than 0.001 |
| Gender vs sleep quality | -- | 0.427 (NS) |

The association between total daily smartphone hours and sleep quality was also significant (chi-square=83.225, p less than 0.001): 93.4% poor sleep in high daily users (6+ hours, n=276) versus 51.6% in lower users (less than 6 hours, n=93). Additionally, high daily users were more likely to have prolonged sleep onset latency: 97.6% of high daily users reported sleep onset latency greater than 30 minutes, compared to 72.6% of lower users (chi-square=47.751, p less than 0.001).

### 3.5 Logistic Regression Analysis

To assess the independent associations of bedtime cessation behavior and total daily screen time with poor sleep quality, binary logistic regression was performed with both variables entered simultaneously (Model 3, Table 3). Bedtime use until sleep onset remained independently and strongly associated with poor sleep quality (OR 15.3, 95% CI 5.7-41.2, p less than 0.001), while total daily screen time was no longer significantly associated (OR 1.8, 95% CI 0.7-4.7, p=0.228). In the fully adjusted model additionally controlling for exam period status, residential setting, and nocturnal phone checking (Model 4), bedtime use until sleep onset remained independently associated with poor sleep quality (OR 10.1, 95% CI 3.2-31.5, p less than 0.001), while total daily screen time remained non-significant (OR 1.5, 95% CI 0.5-4.1, p=0.437).

**Table 3.** Logistic regression models for poor sleep quality as outcome (N=369)

| Variable | Model 1 OR (95% CI) | Model 2 OR (95% CI) | Model 3 OR (95% CI) | Model 4 OR (95% CI) |
| --- | --- | --- | --- | --- |
| Until sleep onset | 24.1 (12.1-48.1)*** | -- | 15.3 (5.7-41.2)*** | 10.1 (3.2-31.5)*** |
| High daily use (6+ hrs) | -- | 13.3 (7.1-24.8)*** | 1.8 (0.7-4.7) NS | 1.5 (0.5-4.1) NS |
| Exam period | -- | -- | -- | 1.0 (0.5-2.1) NS |
| Hostel residence | -- | -- | -- | 1.1 (0.6-2.0) NS |
| Nocturnal phone checks | -- | -- | -- | 2.0 (0.9-4.8) NS |

Model 1: Until sleep alone. Model 2: High daily use alone. Model 3: Both simultaneously. Model 4: Fully adjusted.
\*\*\* $p<0.001$ , NS=not significant. OR=odds ratio, CI=confidence interval.

These findings suggest that the association between total daily screen time and poor sleep quality observed in univariate analyses may be substantially explained by the collinear relationship between heavy phone use and bedtime cessation behavior (correlation r=0.789), rather than representing an independent effect of total use volume.

### 3.6 Moderating Role of Exam Period

Stratified analyses revealed that the association between bedtime smartphone use and sleep quality differed substantially by exam period status. Outside exam periods (n=228), students using smartphones until sleep onset had 99.0% poor sleep quality, compared to 24.2% among those who stopped before sleeping -- a difference of 74.8 percentage points (chi-square=143.912, p less than 0.001). During exam periods (n=141), no significant association was observed between bedtime cessation behavior and sleep quality (71.0% vs 50.9% poor sleep; chi-square=3.167, p=0.075).

### 3.7 Residential Setting

Residential setting was significantly associated with bedtime smartphone behavior. Hostel-dwelling students had the highest prevalence of smartphone use until sleep onset (79.0%, n=169/214), compared to 41.7% in private accommodation (n=25/60) and 23.2% in family-living students (n=22/95) (chi-square=86.609, p less than 0.001).

### 3.8 Academic Performance

Students using smartphones until sleep onset reported substantially worse academic performance: 94.2% reported poor, average, or below-average academic standing, compared to 47.6% of those who ceased use before sleeping (chi-square=102.599, p less than 0.001). Poor sleep quality was associated with academic underperformance: 86.7% of poor sleepers reported below-average or average academic performance compared to 43.3% of good sleepers (chi-square=68.224, p less than 0.001) [18,19]. Low daily study hours (less than 2 hours) were reported by 61.2% (n=226) and were associated with both poor sleep quality (95.9% vs 52.9%; chi-square=89.978, p less than 0.001) and academic underperformance (96.4% vs 53.4%; chi-square=91.102, p less than 0.001). High stress was associated with academic underperformance (92.9% vs 68.8%; chi-square=23.905, p less than 0.001). Gender was not associated with sleep quality (p=0.427).

## 4. Discussion

This cross-sectional survey of 369 medical students in Punjab, Pakistan found that bedtime smartphone use until sleep onset was more strongly associated with poor sleep quality than total daily screen time. Students using smartphones until sleep onset had 95.1% poor sleep quality compared to 44.8% in those who stopped before sleeping. When both variables were entered simultaneously into a logistic regression model, bedtime use until sleep onset remained independently and strongly associated with poor sleep quality (OR 15.3, 95% CI 5.7-41.2, p less than 0.001), while total daily screen time lost statistical significance (OR 1.8, 95% CI 0.7-4.7, p=0.228). These findings suggest that the cessation timing dimension of smartphone use captures the behavioral variance most strongly associated with sleep disruption in this population.

These findings extend the existing literature by identifying cessation timing as a behaviorally specific variable with stronger associations with sleep outcomes than total use volume. Prior studies using composite screen time or addiction scale measures -- including SAS-SV based studies from comparable LMIC populations [8,9,10] -- have not disaggregated the timing of use relative to the sleep-wake cycle from total use volume. Objective wearable sensor data corroborate this finding: Kheirinejad et al. demonstrated that smartphone use specifically in bed significantly worsens sleep latency and awake time, while smartphone use outside of bed does not produce the same effect [13]. The collinear relationship between heavy phone use and bedtime cessation behavior (r=0.789) observed in this sample suggests that total use volume may not independently account for sleep quality when cessation timing is considered.

The exam period moderation finding is a novel contribution that has not been reported in comparable published studies. Outside exam periods, the association between bedtime smartphone use and poor sleep quality was near-total -- 99.0% of students using smartphones until sleep onset reported poor sleep quality, compared to only 24.2% of those who stopped before sleeping. During exam periods, this association was no longer significant (p=0.075), suggesting that exam-related stress may be more strongly associated with sleep quality during those periods, potentially attenuating the observed association with bedtime behavior [14]. This has practical implications for intervention design: digital wellness programs targeting bedtime cessation behavior may be most impactful when implemented and reinforced during non-exam periods.

The residential setting finding adds a structural dimension to the analysis. Students in hostel accommodation were substantially more likely to use smartphones until sleep onset (79.0%) compared to family-living students (23.2%). Students living with family reported lower rates of smartphone use until sleep onset than students residing in hostels or private accommodation. Hostel-level digital wellness programs and structured bedtime environment guidance may therefore be particularly relevant for this subgroup [20].

The observed associations between bedtime smartphone use, poor sleep quality, and academic underperformance suggest a potential pathway linking these three variables. Each association is independently significant: bedtime use until sleep onset is associated with 94.2% academic underperformance versus 47.6% in those who stop before sleeping; poor sleep quality is associated with 86.7% academic underperformance versus 43.3% in good sleepers [3,18,19]. Social media -- the primary smartphone use reported by 36% of respondents -- has itself been independently associated with poor sleep quality in medical students [12], suggesting that the content consumed during bedtime use may compound the timing effect. The additional finding that 61.2% of respondents study less than 2 hours per day -- and that low study hours are associated with both poor sleep quality and academic underperformance -- raises the possibility that smartphone use may be displacing study time alongside its association with sleep disruption, though the cross-sectional design precludes causal conclusions.

The prevalence of poor sleep quality in this sample (75.7%) is higher than rates typically reported in comparable published studies, which range from approximately 40-66% [2,8,9]. This may reflect the extreme bedtime patterns documented here -- 49.1% reported going to bed after 2:00 AM and 87.0% reported sleep onset latency greater than 30 minutes -- suggesting a population with particularly disrupted sleep. These patterns are consistent with the high smartphone use intensity reported: 57.5% reported more than 8 hours of daily use, and 59.9% reported checking their phone every night between midnight and wake time [17].

Several limitations should be acknowledged. The cross-sectional design precludes causal inference; reverse causality is possible -- students with poor sleep may use smartphones more intensively as a consequence of poor sleep rather than as a cause. Sleep quality assessment relied on items informed by PSQI response formats rather than the full validated PSQI scoring protocol with seven component scores, limiting direct comparability with studies using the complete instrument. Smartphone use patterns were self-reported and subject to recall and social desirability bias. The survey did not include a validated smartphone addiction scale such as the SAS-SV, limiting comparability with addiction construct-based studies; this was a deliberate methodological choice, but future studies should consider integrating both approaches. Convenience sampling via social media may over-represent digitally engaged students. The study was conducted in May 2026 -- a single month -- which may not capture seasonal variation in smartphone use or sleep patterns across the academic year. Findings may not generalize to non-digitally engaged students, students outside Punjab, or medical students in other LMICs. The high collinearity between cessation timing and total use volume (r=0.789) warrants caution in interpreting the logistic regression coefficients independently. The wide confidence intervals observed in the logistic regression models reflect imprecision in the effect estimates and should be interpreted with caution.

## 5. Conclusions

Bedtime smartphone use until sleep onset is more strongly associated with poor sleep quality than total daily screen time among medical students in Punjab, Pakistan. This association is more pronounced outside exam periods, structurally patterned by residential setting, and associated with academic underperformance. Medical institutions should consider integrating targeted digital wellness education -- specifically addressing bedtime cessation timing -- into student health programs, with particular attention to hostel-dwelling students where behavioral vulnerability appears greatest [19,20].

## Declarations

### Author Contributions

Mobeen Sajjad: conceptualization, study design, data collection, data analysis, manuscript writing, and final approval.

### Data Availability

The anonymized dataset will be made available upon reasonable request to the corresponding author at.

### AI Use Disclosure

AI tools were used for language refinement during manuscript preparation. All scientific content, interpretations, and references were developed and independently verified by the author.

### Ethics Statement

This study involved anonymous, voluntary online survey participation with no collection of personal identifiers, no clinical intervention, and no patient data. All participants provided informed consent via a mandatory confirmation item. The study qualifies for exemption from formal IRB review under standard criteria for anonymous survey research.

## STROBE Compliance Statement

This manuscript was prepared in accordance with the STROBE statement for cross-sectional studies. A completed STROBE checklist is available upon request from the corresponding author.

